# *U1* snRNA mutation reshapes tumor microenvironment in chronic lymphocytic leukemia: a role for CD44-mediated signaling

**DOI:** 10.1101/2025.03.31.25324954

**Authors:** Sara López-Tamargo, Javier Fernández-Mateos, Pablo Bousquets-Muñoz, Laura Llaó-Cid, Ferran Nadeu, Ares M. Farran, Cristina Olivar-Fernández, Ana de la Fuente-González, Andrea Aran, Roberto Martínez-Soler, Europa Azucena González, Manel Juan, José I. Martín-Subero, Dolors Colomer, Elías Campo, Ana Gutiérrez-Fernández, Xose S Puente

## Abstract

Chronic lymphocytic leukemia (CLL) is characterized by the accumulation of monoclonal mature B lymphocytes in peripheral blood, bone marrow, lymphoid tissues, and extranodal sites. Genes involved in RNA splicing such as *SF3B1* and *U1* are frequently mutated in CLL, leading to altered splicing and generation of tumor neoepitopes. To study the impact of these mutations on the tumor microenvironment (TME), we have developed a comprehensive single-cell atlas of unmutated CLL encompassing 26 bone marrow and lymph node tumor samples from 23 U-CLL patients with mutations in *U1* (n=7), *SF3B1* (n=8), or without mutations in splicing genes (n=10). We observed high intra-tumor heterogeneity, discerning 12 transcriptional programs, one linked to the *U1* g.A3>C mutation and characterized by NFKB hyperactivation. T cell and NK compartments exhibited site- and mutation-specific enrichment, with increased CD4^+^ regulatory cells (Treg) and CD8^+^ exhausted cells in lymph nodes, while *U1*-mutant tumors showed increased CD8^+^ cytotoxic activity, with a predominance of effector-like CD8^+^ cells. Single-cell T cell receptor sequencing revealed clonotype expansion in *U1*-mutated tumors, particularly in CD8^+^ effector and exhausted cells, suggesting a neoantigen-driven immune response. Cell-to-cell interaction analysis identified CD44 as a key mediator in *U1*-mutated tumors, showing pro-B survival interactions as those involving MIF-CD44-CD74. Furthermore, interactions between CD80 on CLL cells and CTLA4 on Tregs and CD8^+^ exhausted were upregulated, reflecting an immunosuppressive phenotype associated with *U1* mutated CLL. These findings highlight the complex interplay between mutations in CLL and the TME, offering novel avenues for alternative therapeutic strategies for U-CLL with mutations in *U1*.

## Introduction

Chronic lymphocytic leukemia (CLL), the most frequent leukemia in adults, is characterized by the progressive accumulation of monoclonal mature B lymphocytes in peripheral blood, bone marrow, lymphoid tissues, and extranodal sites^1^. The biological and clinical evolution of the disease exhibits a heterogeneous course, with patients manifesting either an indolent disease or an aggressive progression. CLL is classically divided into two main subgroups based on the presence or absence of acquired somatic mutations in the immunoglobulin heavy chain gene (IGHV) of the B-cell receptor (BCR) expressed by leukemic B cells^2,3^.

Over the last 15 years, genome sequencing studies have provided a comprehensive characterization of genetic and genomic alterations in CLL in more than 1,000 tumors ^4–7^. Globally, around 100 putative driver genes and 100 recurrent structural variants were identified in CLL^8^, but only a few genes are mutated in more than 5% of tumors at diagnosis, including *SF3B1*, *NOTCH1*, *ATM* or *TP53*, followed by a long tail of genes mutated at lower frequencies^8^. Despite the diverse number of mutated genes in CLL, most are clustered in a small number of cellular pathways^9^. Mutations in genes involved in RNA splicing are of particular interest due to both their high frequency, the association with specific stereotypes and epigenetic subgroups^10^, and their pleiotropic effect on multiple genes, causing alterations in the splicing of oncogenes and tumor suppressors^11,12^.

Notably, the most frequently mutated gene, *SF3B1*, encodes a crucial protein for the binding of the spliceosomal *U2* snRNP to the branch point proximal to the 3’ splicing site. Recurrent somatic point mutations in this gene perturb the splicing machinery, leading to differential splicing junctions and enhanced expression of truncated mRNAs in CLL cases^13,14^. Recently, a recurrent mutation at the third base of the *U1* small nuclear RNA (snRNA) has been identified in different tumors, including CLL. This snRNA forms part of the spliceosome and is involved in the recognition of the 5’ splice-site of RNAs. A g.3A>G mutation in *U1* is present in 19% cases of medulloblastoma^15^, while a g.3A>C substitution occurs in 4% of CLL cases, particularly associated with unmutated-IGHV CLL (U-CLL)^12^.

Due to the relevance of *U1* snRNA in the recognition of the splicing donor site, *U1*-mutant tumors show a major alteration in RNA splicing, with an excess of 5’ cryptic splicing events. The generation of numerous aberrantly spliced transcripts might lead to the synthesis of abnormal proteins, resulting in the generation of tumor neoepitopes that could be recognized by the immune system. However, despite the potential effect of splicing mutations on the activation of an immune response, tumors with mutations in these splicing genes are associated with a more aggressive disease and poor outcome, both for *U1*^12,15^ and *SF3B1* ^13^. These results suggest that mutations in splicing genes might result in a disruption of the crosstalk between CLL tumor cells and the tumor microenvironment (TME). This is especially important in CLL biology, as malignant cells proliferate almost exclusively in specific niches such as bone marrow or lymph nodes^16^, underscoring a high dependence on TME.

In this study, we conducted an in-depth profiling of CLL using single-cell sequencing to analyze tumor cells and the TME landscape in the context of splicing dysregulation. This analysis revealed discernible alterations both in CLL cells as well as in the cellular composition of the TME, with the identification of differential enrichment of specific cellular signals in tumors, depending on their location and the presence of splicing driver mutations. Cell-to-cell interactions also revealed the activation of specific molecular pathways, particularly in *U1*-mutated samples, which could provide novel therapeutic targets in U-CLL.

## Results

### Generation of a single-cell atlas for U-CLL

Given the higher frequency of mutations in splicing genes observed in U-CLL compared to M-CLL cases, we analyzed 26 tumor samples from 23 U-CLL patients with mutations in either *U1* (n=7), *SF3B1* (n=8) or without mutations in splicing genes (n=11)(**Figure 1A-B**). According to its tissue, 15 samples were obtained from bone marrow, while 11 from lymph node. Clinical features and single-cell metrics are summarized in **Supplemental Table 1** and **Supplemental Figure 1A-C**, while the mutational landscape is shown in **Figure 1B**. Qualified data from 160,317 single cells were finally considered for downstream analysis, 61% of them corresponding to tumor B-cells, although tumor purity was variable among samples (74-95%). The most abundant compartment within the TME was CD8^+^ T (8.2%), followed by erythroblasts (6.4%), and CD4^+^ T cells (5.2%) (**Figure 1C-D**). Clusters were annotated based on known markers both in RNA and surface protein expression (**Supplemental Figure 1D-E)**.

**Figure 1.**
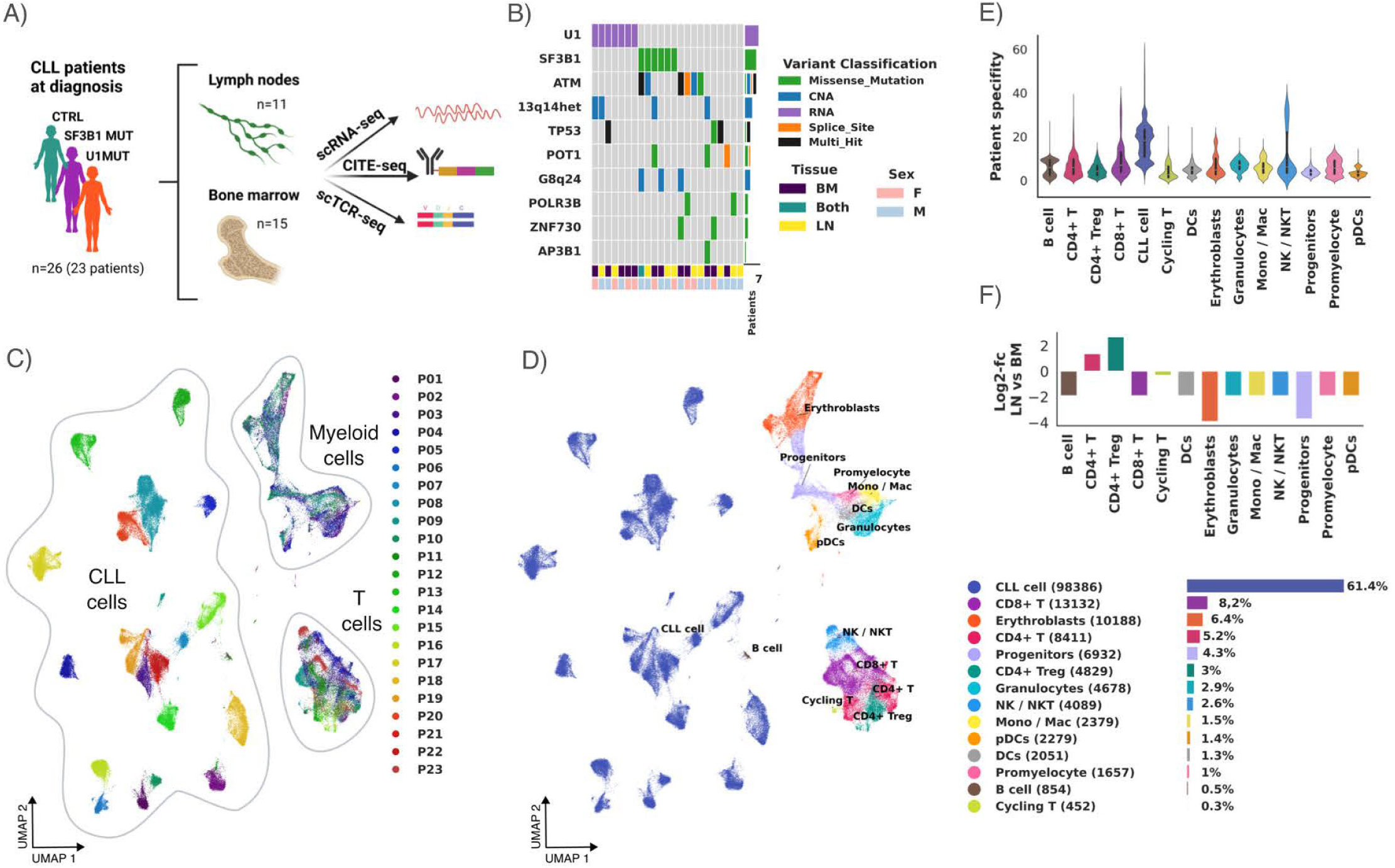
Unmutated CLL single-cell atlas. (A) Overview of sample collection and sequencing strategy. (B) Oncoplot depicting the main driver genes and copy number alterations present in each analyzed patient. (C) UMAP projection of scRNAseq data colored by patient, showing the distribution of CLL, myeloid and T cells. (D) UMAP projection colored by major cell types, accompanied by a legend showing the relative and absolute frequencies. (E) Patient specificity score for each cell type. (F) Enrichment of cell type composition expressed in log2FC for lymph node compared to bone marrow.

UMAP projection showed that tumor cells clustered by patient, while non-tumor cells (T cells and myeloid cells) clustered together with independence of the patient of origin (**Figure 1C-D**). A patient specificity score based on cell neighboring showed that tumor B-cells exhibited a higher patient specificity (*p*<2 x10^-16^) than other cell types (**Figure 1E**), possibly reflecting the tumor particular mutational profile and/or epigenetic alterations, while the remaining TME components were more transcriptional homogeneous at the same level of integration (**Figure 1C-E**).

Despite the overall similarity of TME components between samples, there were major differences in cell populations based primarily on the tissue of origin. Overall, lymph node-derived TME was characterized by an increase in CD4^+^ Treg (log2FC=2.55) and CD4^+^ T cells (log2FC=1.19) compared to bone marrow (**Figure 1F** and **Supplemental Table 2**). In contrast, bone marrow-derived TME showed a significant enrichment in hematopoietic precursors such as common myeloid progenitors-CMP (log2FC=2.00) and erythroblasts (log2FC=3.87) when compared to lymph node samples (**Figure 1F** and **Supplemental Table 2**).

### NMF uncovers transcriptionally heterogeneous tumor cell subpopulations

To discern latent transcriptional programs within the tumoral fraction, we decomposed gene expression using Bayesian Non-negative Matrix Factorization (NMF), obtaining a total of 12 gene signatures (**Supplemental Figure 2A-D**). Top representative genes in each signature were not only correlated by their expression patterns, but also appeared to exhibit underlying biological significance, as many of these genes were functionally related (**Figure 2A** and **Supplemental Table 3**). Some of these signatures seemed to be similar to others previously described, including a histone-related signature corresponding to S8^17^ and a glycolysis-related signature, also called *activated CLL signature*, represented by S12^18^.

**Figure 2.**
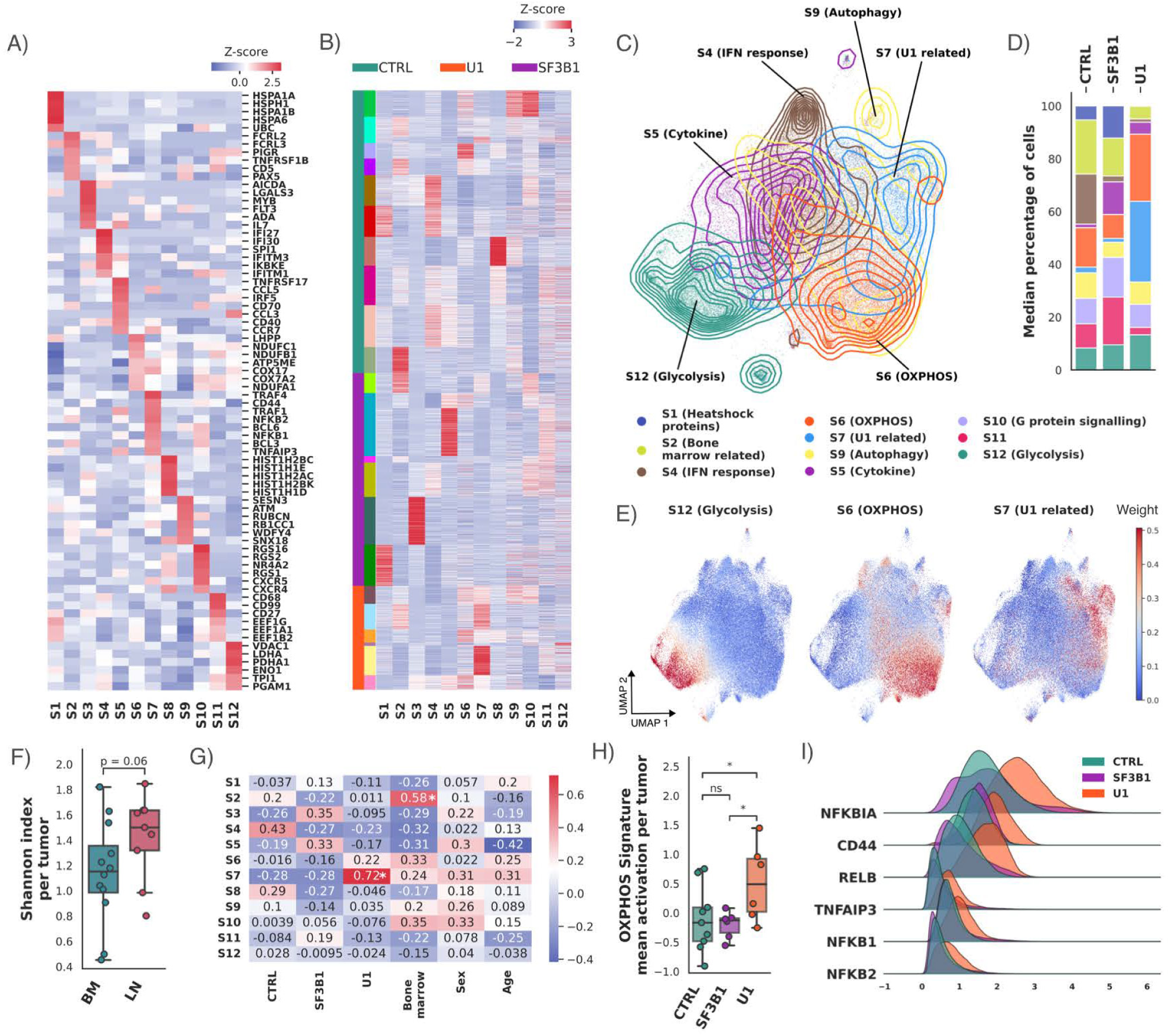
Intratumoral heterogeneity of tumor CLL cells. (A) Heatmap showing selected top genes used to annotate the NMF programs and their relative importance, normalized by z-score of their weight. (B) Heatmap showing relative importance of NMF programs in each cell, ordered by condition and patient. The first legend clusters tumor cells by condition, and the second one cluster cells by patient. (C) Contour plot of gaussian kernel density estimations of selected signatures projected onto the UMAP plot. (D) Condition-specific composition of signatures. Each cell was classified based on its maximum signature and the percentage of maximums in each patient was used to calculate the media per condition. (E) UMAP plot of tumor cells colored by the weight of glycolysis, OXPHOS and *U1*-related signatures, respectively. (F) Intratumor transcriptional diversity estimated by Shannon index per patient grouped by tissue. Shannon index was calculated in each patient separately considering each cell as a specimen and identified by its maximum signature. The p-value displayed corresponds to a Mann-Whitney pairwise comparison. (G) Heatmap with pairwise Pearson correlation between NMF signatures and study variables. *, *p*<0.05. (H) OXPHOS signature mean activation per tumor grouped by condition. The p-value displayed corresponds to a Mann-Whitney pairwise comparison. (I) Ridge plot showing gene expression distribution grouped by condition. Selected genes are both top 250 genes of S7 signature and differentially upregulated in *U1* tumor cells.

Most transcriptional programs were shared among different tumors regardless of tissue and condition (**Supplemental Figure 2E**), with the exception of two signatures, S3 characterized by elevated levels of deaminases and exclusive to patient P17, and S8, associated with histones and specific to patient P16 (**Figure 2B**). Those two patient-specific signatures, both from the control group, were not considered for further analyses.

The different transcriptional programs were distributed along the UMAP clustering. As an example, the two main metabolism-related signatures, S12 characterized by glycolysis, and S6 enriched in oxidative phosphorylation (OXPHOS) programs, were opposite displayed in the plot and showed a mutually exclusive activation (**Figure 2C-E**). We observed a greater transcriptional diversification of CLL cells from lymph node compared to bone marrow using the alpha diversity analysis based on Shannon index (*p*=0.063), which can be attributable to the possible different influence of these two TME on the transcriptional programs of the tumor cells (**Figure 2F**).

In order to determine whether any of these transcriptional programs was associated with the tissue of origin or mutational status of the tumors, we performed correlation and multivariable regression analyses (**Supplemental Table 4**). We observed that signature S2, characterized by the expression of genes such as *PAX5*, *CD5* or *TNFRSF1B*, involved in the development of hematopoietic cells, was associated with bone marrow-derived samples (*p*=0.0038) (**Figure 2G**). Interestingly, the highest correlation was found between the S7 transcriptional program and the presence of the *U1* g.3A>C mutation (*p*=0.0049), while the S6 program, characterized by the overexpression of mitochondrial respiratory chain (OXPHOS) genes, also showed an increase in *U1*-mutated tumors when compared to the control and *SF3B1* groups (*p*=0.033) (**Figure 2H**). Conversely, no statistically significant association was found between any of the identified transcriptional programs and the presence of mutations in *SF3B1*. Together, these data suggest that both the tumor genomic landscape and the anatomical location have an effect on the gene expression program of the tumor compartment.

### S7 signature and NFKB signaling associated genes are upregulated in U1-tumors

To further understand the biological relevance of the S7 signature, we performed pathway enrichment analysis, with the finding of nuclear factor of kappa B (NFKB) signaling as the most significantly enriched pathway among genes within the S7 transcriptional program (**Supplemental Figure 2F**). Although activation of NFKB signaling in CLL has been associated with the lymph node TME^19^, this pattern was observed in all *U1*-mutated CLL samples, most of them bone marrow-derived, showing that this activation was independent of the tissue of origin. To understand the directionality of these changes, we inferred the weight of transcription factor networks with a univariate linear model. An increase in both NFKB (*p*=0.016) and NFKBIB (*p*=0.028) was identified, reflecting a global activation of this pathway (**Supplemental Figure 2G-H**).

To identify individual genes with altered expression in the S7 transcriptional program, a differential expression pseudobulk analysis was performed, showing an enrichment in genes related to maturation of ribosomal RNA and ribosome biogenesis (**Supplemental Figure 2I**). As expected, some of the most significant genes belonged to the NFKB pathway including *NFKBIA* (*p*=5.87×10^-6^, log2FC=1.04), *CD44* (*p*=0.02, log2FC=0.61), *RELB* (*p*=5.21×10^-6^, log2FC=0.82), *TNFAIP3* (*p*=7.62×10^-6^, log2FC=1.08), *NFKB1* (*p*=7.59×10^-6^, log2FC=0.58) and *NFKB2* (*p*=*1*.18×10^-5^, log2FC=0.64), among others (**Figure 2I** and **Supplemental Table 5**). This result provides a connection between the alterations in splicing caused by the *U1* g.3A>C mutation and the NFKB signaling pathway. In this regard, it is worth noting that one of the top genes contributing to signature S7 was *CD44*, previously identified as one of the genes with most splicing changes induced by the expression of the *U1* g.3A>C mutation^12^.

### U1 mutated tumors exhibit an increased CD8^+^ T cytotoxic activity

To further characterize the TME component, we proceeded to identify subgroups within the T and NK compartments. We annotated a total of 30,270 cells (18.88%). CD4^+^ cells were divided into 3 groups: effector/memory (15.60%), Treg (11.40%) and naïve/central memory (naïve/Tcm) (7.85%), while CD8^+^ cells were subclassified into: effector/memory (33.75%), exhausted (TEX) (11.69%), and naïve/ Tcm (4.65%). NK and NKT cells did not cluster separately and were included under the NK/NKT label (13.40%) (**Figure 3A** and **Supplemental Figure 3A-B**).

**Figure 3.**
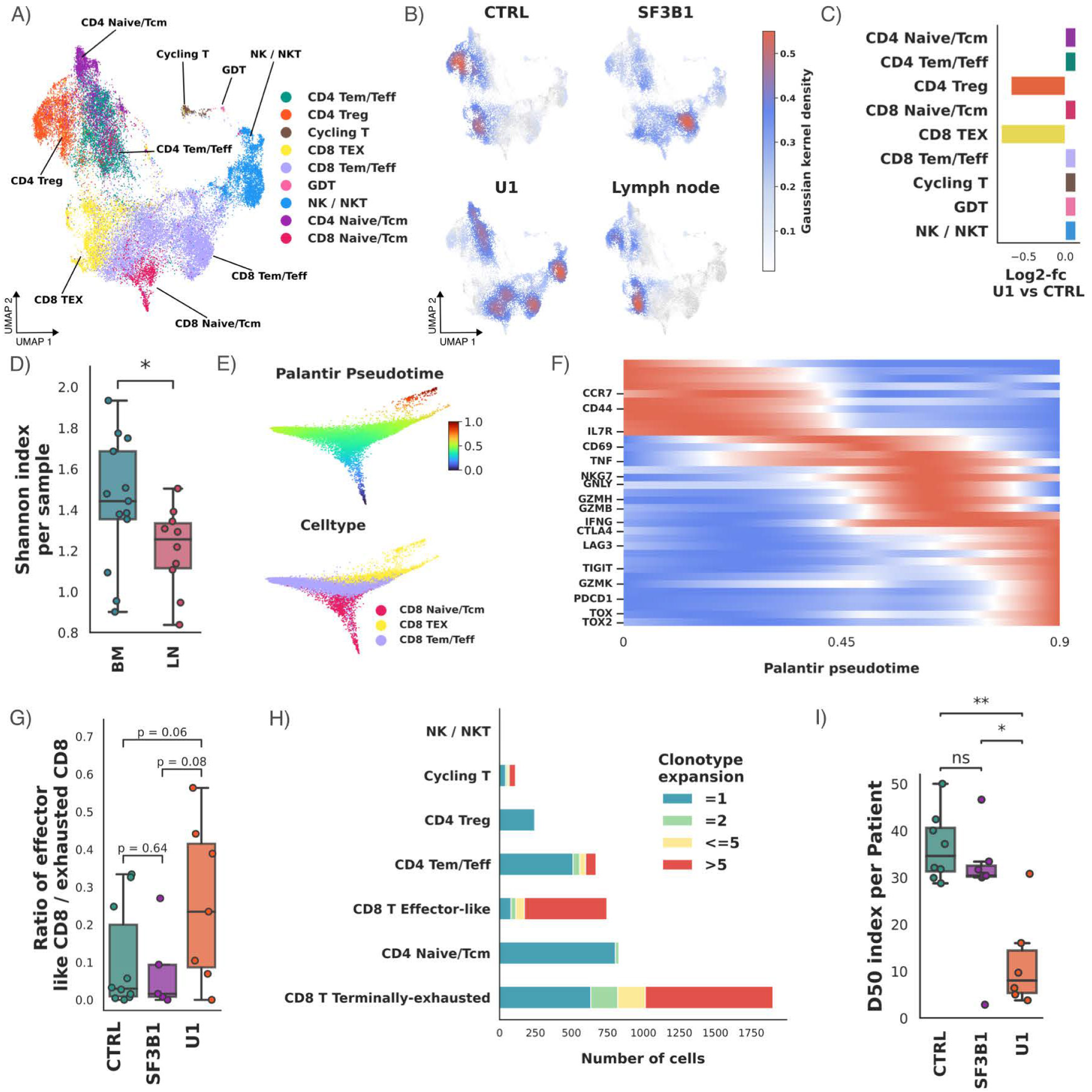
Profiling of T/NK cell tumor microenvironment. (A) UMAP projection of the T and NK compartment profiled by scRNAseq, colored by manually annotated cell subtypes. (B) Gaussian Kernel density estimation of T/NK subtypes projected onto the UMAP plot in Control, *SF3B1*, *U1* condition and lymph node, respectively. (C) *U1* condition specific enrichment of cell type composition expressed in log2FC compared to the control. (D) Intra-tumor population diversity estimated by Shannon index per patient grouped by tissue. The p-value displayed corresponds to a Mann-Whitney pairwise comparison. *, *p*<0.05. (E) Palantir trajectory analysis of CD8+ T cell subset colored by pseudotime and T subtype annotation, respectively. (F) Gene expression dynamics of CD8+T exhausted branch across pseudotime. Selected genes are labeled. (G) Ratio of effector-like CD8+ T to exhausted CD8+ T per patient grouped by condition. Each cell was assigned to a specific pseudotime differentiation branch based on their fate probabilities using Palantir algorithm. The percentage of each intra-tumoral CD8+ T state was calculated for each patient. The p-value displayed corresponds to a Mann-Whitney pairwise comparison. (H) Clonotype expansion per cell subtype in the *U1* condition. Clonotype expansion was transformed into a categorical variable based on the number of cells detected inside a clonotype. (I) Diversity 50 (D50) index of TCR repertoires per patient grouped by condition in CD8+ compartment. ***, *p*<0.001. The p-value displayed corresponds to a Mann-Whitney pairwise comparison.

T and NK subclusters distribution varied across different contexts, exhibiting a site and condition-specific enrichment (**Figure 3B-C**). The highest changes were observed between tissues, with CD4^+^ NK/NKT and CD8^+^ Tem/Teff significantly enriched in bone-marrow-derived samples when compared to lymph node-derived (log2FC=1.93 and 1.90, respectively) (**Supplemental Figure 3C-D** and **Supplemental Table 6**). Conversely, the CD8^+^ TEX population was more expanded in lymph node (log2FC=2.45). Comparison of the mutational status revealed significant differences between samples derived from *U1*-mutated tumors and the control group, with a marked reduction of CD4^+^ Treg (log2=-0.85) and CD8^+^ TEX (log2=-0.80) (**Figure 3C** and **Supplemental Table 6**), independent of tissue-specific effect. However, no statistically significant changes were observed when comparing *SF3B1* group versus control.

To measure intra-sample diversity, we estimated Shannon index sample-wise based on T cell subpopulation frequencies. We did not find significant changes associated with the mutational status, but we observed differences correlated to the tissue of origin, with a higher diversity in samples derived from bone marrow (*p*=0.025) (**Figure 3D**).

Due to the observed differences in CD8^+^ T cells in *U1*-mutated samples, we computed the differentiation trajectories of CD8^+^ T cells using Palantir pseudotime. This revealed two terminal traits: the trajectory reaching the end of pseudotime was defined as terminally exhausted CD8^+^, characterized by the loss of naïve (*CCR7* and *CD44*) and cytotoxic markers (*NKG7*, *GZMH*, *GNLY* and *GZMB*), alongside the accumulation of exhaustion markers (*PDCD1*, *TIGIT*, *TOX*, *TOX2*, *CTLA4* and *GZMK*) (**Figure 3E-F**), whereas the other branch distinguished effector-like CD8^+^ T cells accumulating the mark of genes such as *NKG7, IFNG, KLRD1, GZMB, GZMH, GNLY,* and *CTSW* (**Supplemental Figure 3E-F**).

To understand the relationship between the mutational profile of the tumor and the CD8^+^ T cell status, individual cells were assigned to one of the branches based on the most probable predicted fate, resulting in 23.4% of effector-like T cells and 76.6% of TEX. We observed a difference when effector/exhausted ratio was measured, with a predominance of cytotoxic phenotype in *U1* tumors (*p*=0.059) when compared to control or *SF3B1* mutated samples (**Figure 3G**).

### U1-mutated tumors show a higher clonotype expansion of CD8^+^ T cells

The presence of the *U1* g.3A>C mutation triggers widespread splicing-related alterations, what might contribute to the generation of neoepitopes that can induce an immune response. To determine whether the presence of the *U1* g.3A>C mutation could have an impact on TCR clonality, we performed single-cell TCR sequencing in all samples. We were able to identify 10,300 unique TCR beta (TCRβ) sequences in 14,897 cells for which a TCR sequence was obtained (**Supplemental Figure 4A)**. The TCR immunorepertoire was variable across samples, and there were no clonotypes shared among patients. There was one case (P08) for which two biopsies were available from different tissues (bone marrow and lymph node). Analysis of the TCRs in these samples revealed the presence of common clonotypes shared between different tissues (N=12), revealing a remarkable similar clonal composition and suggesting that these cells might constitute tumor-reactive T cells. CD8^+^ TEX was the compartment with the most presence of shared clonotypes (60%), followed by CD8^+^ Tem/Teff and cycling T.

Whereas some TCR repertoires showed minimal clonal expansion, others were predominantly composed of a small number of dominant T cell clones, with high inter-patient variability (**Supplemental Figure 4B-C**). Interestingly, we observed a non-significant trend towards clonal expansion in *U1*-mutated samples, estimated by the normalized Shannon Entropy and Diversity 50 (D50) (*p*=0.1). To determine the cell types in which this expansion occurred in *U1*-mutant cases, clonotype frequencies were quantified in each T cell subtype. This analysis revealed that the expansion predominantly took place in CD8^+^ T cells, with most effector-like CD8^+^ cells grouped in clonotypes, followed by terminally exhausted CD8^+^ T cells (**Figure 3H** and **Supplemental Figure 4D**). When we performed the diversity analysis only to CD8^+^ cells, we observed a higher significant clonal expansion in *U1*-mutated samples when compared to both control and *SF3B1* groups (*p*=0.001) (**Figure 3I**). The *SF3B1*-mutated group also exhibited an expansion of clonotypes, primarily within the CD8^+^ compartment; however, the extent of this expansion was not significantly higher than that observed in control samples (**Supplemental Figure 4D-F**). Notably, there were shared clonotypes between different CD8^+^ populations. In particular, in *U1*-derived samples, 42% of the expanded CD8^+^ clonotypes with more than five cells were shared between CD8^+^ effector and exhausted lineages (**Supplemental Figure 4G**). Multiple regression analysis also revealed that tissue had an effect on the CD8^+^ clonal expansion, being higher in bone marrow (**Supplemental Figure 4H**). These data suggest an antigen-driven expansion of specific CD8^+^ populations that later followed their terminal differentiation as effector or exhausted CD8^+^ cells.

### CLL cells show a higher auto stimulation through CD44 in U1–mutated tumors

To gain further insights into the regulatory relationship between T cell subclusters and tumor B-cells, immune-related ligand-receptor (L-R) pair interactions were decomposed into eleven different latent factors using Tensor Component Analysis (TCA) (**Supplemental Figure 5A** and **Supplemental Table 7**). The analysis of these factors revealed differential patterns associated with the tissue of origin and mutation type (**Supplemental Table 8**). Thus, five factors were linked to the tissue of origin, including factors F7 (*p*=9.6×10^-4^), F8 (*p*=1.5×10^-2^) and F10 (*p*=1.06×10^-5^) associated with bone marrow, while factors F4 (*p*=2.6×10^-2^) and F11 (*p*=6.5×10^-6^) were associated with lymph node (**Supplemental Figure 5B-E**). Factors F10 and F11 were the ones with the highest effect, and top-rank interactions for factor F11 included cytokines such as CXCL13, CXCL17, CCL22 and CCL19 (**Supplemental Table 9**), commonly up-regulated in lymph node^20,21^. Additionally, factor F1 was associated with the presence of the *U1* mutation (*p*=3.43×10^-3^) (**Supplemental Figure 5E** and **Supplemental Table 8**). It was also observed a non-significant (*p*=0.07) positive correlation between factor F2 and *U1*-mutated condition.

The observed cell-to-cell interactions for most differential factors did not show a predominant cell type monopolizing the cross-talk between TME cells and tumor B-cells, with a balanced bidirectional signaling (**Supplementary Table 10**). However, factor F1 exhibited a predominant polarity from the TME towards tumor B-cells (**Figure 4A**), while factor F2 showed an opposite behavior, with directionality from CLL cells towards TME compartment (**Figure 4B**). Both factors manifested homotypic interactions between tumor B-cells. Examining the top-rank ligand-receptors in the *U1-* associated factors, we found several statistically significant interactions. First, we noticed an enrichment of connections between *CD44* as the common receptor in factor F1, with extracellular matrix proteins including collagens *COL1A1* (*p*=4.67×10^-2^), *COL6A1* (*p*=3.63×10^-2^) and *COL9A2* (*p*=3.63×10^-2^), laminins *LAMB1* (*p*=1.12×10^-2^) and *LAMB3* (*p*=2.37×10^-3^), and other proteins such as *MMP9* (*p*=1.56×10^-2^), *HGF* (*p*=2.37×10^-3^), *MIF* (*p*=1.56×10^-2^) and *HAS2* (*p*=4.99×10^-4^) (**Figure 4C** and **Supplemental Figure 5F**). These connections might be a result of *CD44* upregulation in *U1*-mutated CLL cells when compared to unmutated ones. The relationship between the *U1* mutation and *CD44* overexpression was independently validated across a cohort of different CLL patients^8^ by bulk RNAseq (N=93, *p*=4.6×10^-4^) (**Supplemental Figure 5G** and **Supplementary Table 11)**. Nonetheless, this communication could also be due to the effect of the *U1* g.3A>C mutation on the splicing of *CD44*^12^. In fact, analysis of *CD44* splicing from single cell sequencing data, as well as through long-read sequencing, confirmed that while control and *SF3B1*-mutated CLL cells express the standard isoform of *CD44* (CD44s), *U1*-mutated CLL cells show specific inclusion of variant exons generating the variant isoforms of *CD44* (CD44v) including the one expressing exon 11 (CD44v6) (**Figure 4D**).

**Figure 4.**
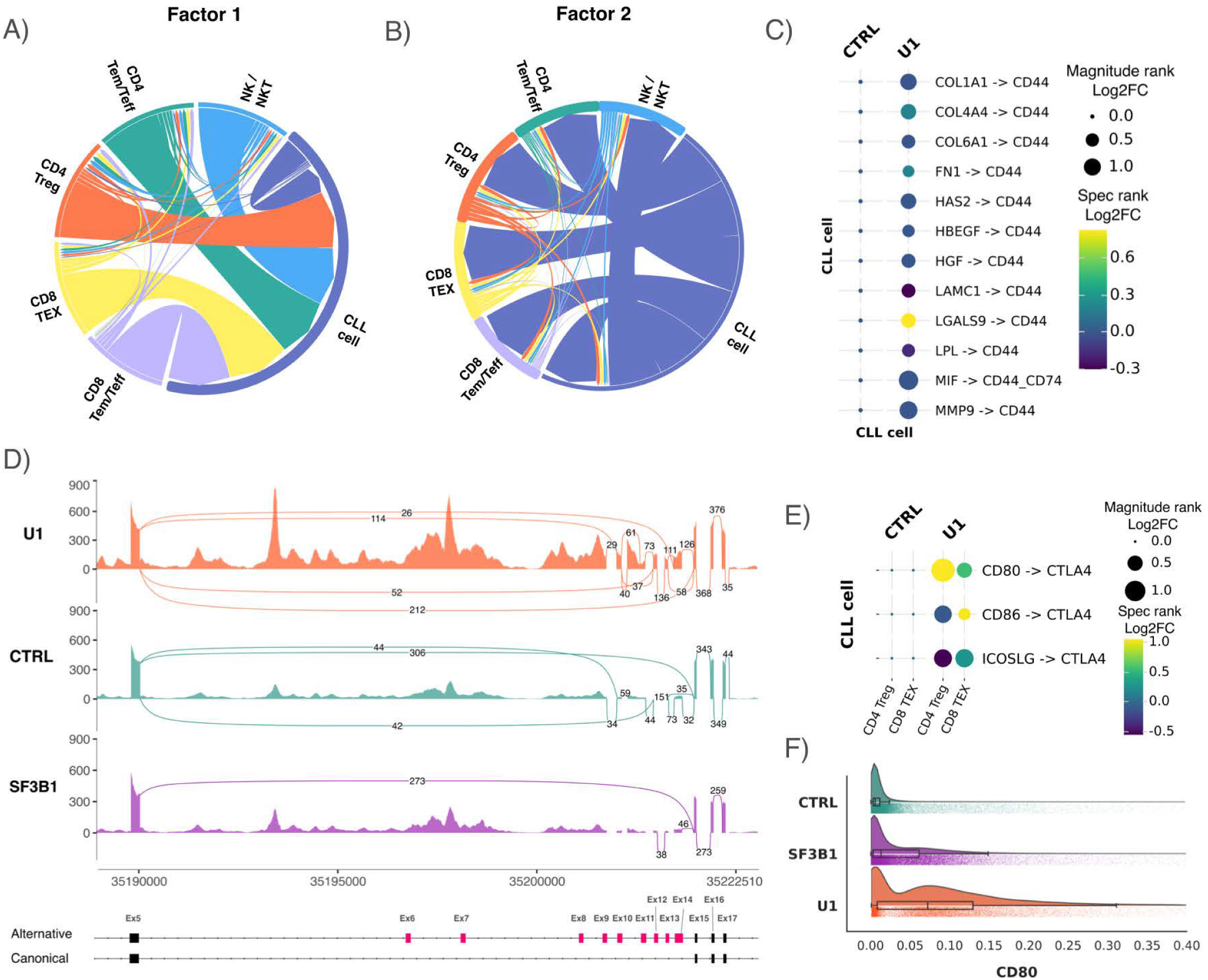
Cell to cell interaction profiling. (A-B) Directionality of factor F1 (A) and factor F2 (B) specific interactions colored by cell type. (C) Dot plot showing top ligand-receptor interactions in factor F1. Both the magnitude and specificity are expressed as the magnitude of change compared to the control, computed as the negative logarithm (-log2) of the ratio of *U1* score to Control score. The source of the interaction is represented on the Y-axis, while the target of the interaction is shown on the X-axis. (D) Sashimi plot showing the *CD44* splicing in tumor cells grouped by condition, indicating the structure of the standard and variant exons on the bottom panel. (E) Dot plot showing top ligand-receptor interactions in factor F2 as in C. (F) *CD80* expression distribution in CLL cells grouped by condition.

The analysis of the top signals contributing to factor 2, revealed the presence of interactions involving *CTLA4* with *CD80*, *CD86* and ICOSLG. In fact, the interaction with the major magnitude was between *CD80* in CLL cells and *CTLA4* in CD4^+^ Treg (*p*=2.09×10^-2^) and CD8^+^ TEX (*p*=3.18×10^-2^) cells (**Figure 4E** and **Supplemental Figure 5H-I**). In addition to the changes in the predicted interactions in factor F2, we observed a significant increase in the expression of *CD80* in tumor cells in the *U1* condition (log2FC=2.66, *p*=2.1×10^-3^) (**Figure 4F** and **Supplemental Table 5**).

## Discussion

CLL constitutes a useful model to understand the complex interplay between tumor cells and their environment, as CLL cells proliferate almost exclusively in the bone marrow or lymph node, reflecting their dependence on prosurvival signals from the TME^22^. However, the extent to which tumor B cells shape their microenvironment, and conversely, remains an open question. In this study, we provide the first comprehensive single-cell atlas of the IGHV-unmutated CLL and its TME, identifying key mechanisms that influence tumor behavior caused by mutation of the splicing factor *U1*.

In line with previous observations in other tumors, our data revealed high intra-tumor heterogeneity^23^, where 12 transcriptional programs with underlying biological significance were characterized, including a glycolysis and an OXPHOS signatures previously identified as relevant in CLL progression^18,24,25^. We showed that one of the transcriptional programs detected in CLL cells is clearly associated with bone marrow, corroborating previously described tissue differences^26^. Interestingly, a transcriptional program (S7) was found to be strongly associated with the presence of the *U1* g.3A>C mutation, and characterized by the hyperactivation of the NFKB pathway regardless of tissue type. High levels of NFKB signaling may contribute to the more aggressive phenotype shown in patients with the *U1* g.3A>C mutation, characterized by a shorter time to first treatment^12^. No transcriptional program was associated with *SF3B1* mutations, possibly due to the limited sample size, the presence of some samples with subclonal mutations, and the relatively modest impact of these mutations on splicing.

Our analyses revealed a compositionally diverse TME, with an enrichment of particular cell types linked to either the anatomical location or mutational status. The finding of an enrichment of CD8^+^ Tem/Teff and NK/NKT cells in bone-marrow, points to a repressed immune response in lymph node relative to bone marrow. This pattern suggests an ineffective host antitumor immune response, similar to that found in classical Hodgkin lymphoma (cHL)^27^. Furthermore, this effect seems to be more pronounced in *U1*-mutated samples, where the reduction in CD4^+^ Treg and CD8^+^ TEX appears to exceed that observed due to the tissue of origin.

The integration of comprehensive T cell trajectories and single-cell resolution TCR immunorepertoire allowed us to classify the clonotype expansion in these samples. We found a clonal expansion, predominantly in CD8^+^ cells, and specifically concentrated within the effector-like CD8^+^ T compartment in *U1*-mutated samples. The fact that most expanded clonotypes were shared between effector and exhausted T cells, suggests a progression towards dysfunctional CD8^+^ T cells, typical of a sustained antigen-induced chronic inflammation such as the one observed during viral infections or cancer^28,29^. We hypothesize that the higher prevalence of expanded clonotypes in *U1*-mutated samples might result from an immune response targeting tumor-neoepitopes generated by aberrant splicing events. Nonetheless, further studies will be required to characterize *U1* g.3A>C-induced neoepitopes, as they could represent promising targets for immunotherapy. Conversely, while mutations in *SF3B1* induce the generation of neoepitopes that lead to the expansion of CD8^+^ T cells in uveal melanoma^30,31^, clonotype expansion was not significantly higher in *SF3B1*-mutated CLL samples than controls. This result suggests that tissue may influence either the type or quantity of neoepitopes generated, as well as their efficacy in eliciting an immune response.

In this regard, one of the paradigms of the *U1* g.3A>C mutation is its ability to induce a cytotoxic response while simultaneously being associated with a poorer prognosis in CLL^12^. The analysis of cell-to-cell interactions revealed the presence of two factors linked to the *U1* mutation, highlighting its broad impact on the TME (**Figure 5**). Of special relevance is the identification of a CTLA-4 stimulation of CD4^+^ Treg cells through CD80/CD86/ICOSLG in CLL cells. This interaction, partially enhanced by the increased expression of *CD80* in *U1*-mutated CLL cells, might contribute to enhancing antigen-specific adaptive Tregs, favoring an immunosuppressive tumor response^32^. Moreover, CTLA-4 is able to deplete CD80/CD86 from antigen-presenting cells, via trans-endocytosis, thereby reducing their capacity to provide co-stimulatory signals and dampening the activation of effector T cells^33,34^. Together, these interactions might contribute to maintain an immunosuppressive phenotype towards *U1*-mutated CLL despite the likely presence of neoepitopes capable of triggering TCR recognition.

**Figure 5.**
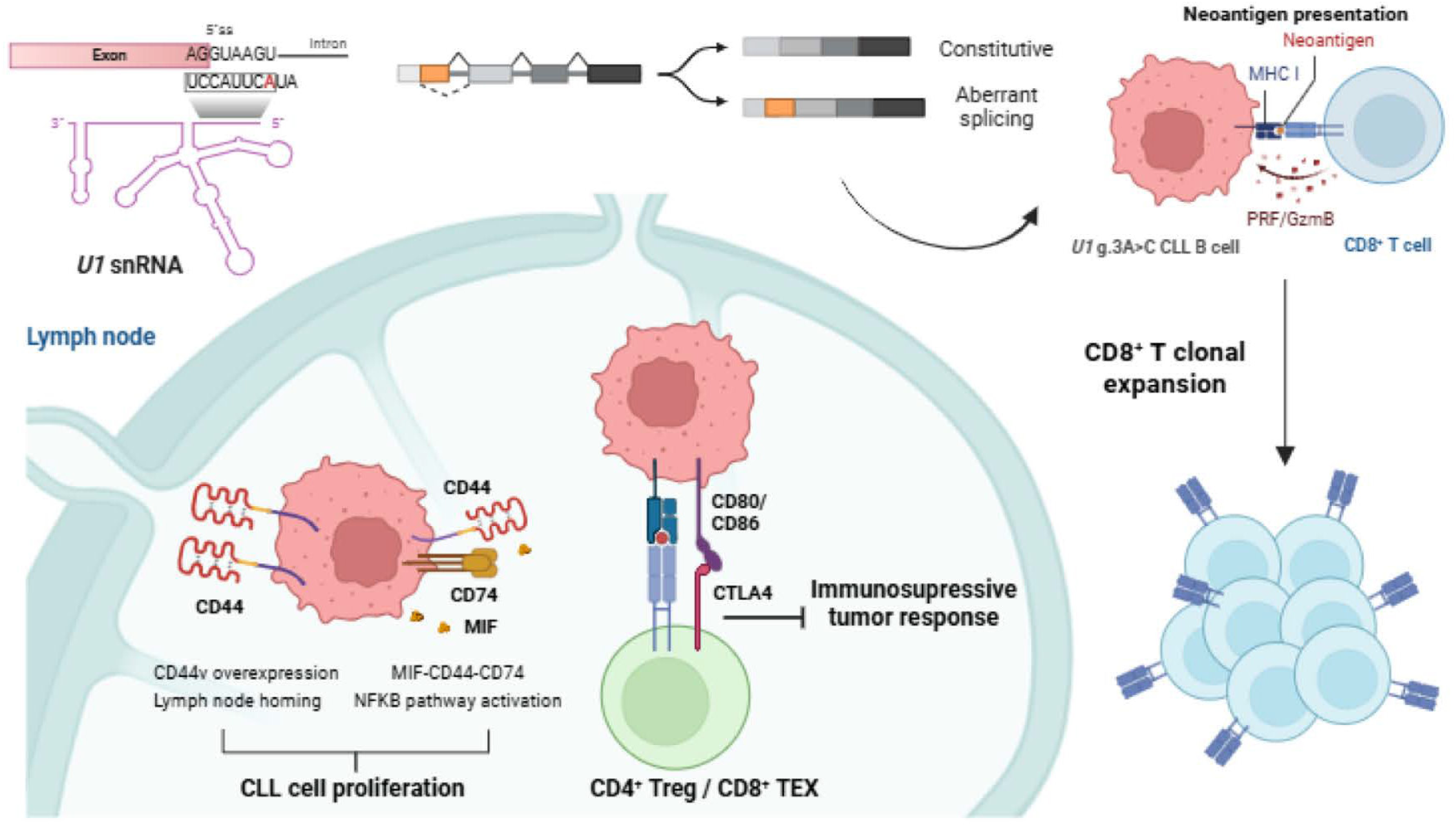
Impact of the *U1* g.3A>C mutation on CLL and TME. Aberrant splicing caused by the U1 g.3A>C mutation results in the generation of neoepitopes and clonal expansion of CD8^+^ T cells. Higher expression of CD44v facilitates homing and together with CD74, increased activation of the NFKB pathway in CLL tumor cells, resulting in higher proliferation and survival of tumor cells. Higher expression of CD80/CD86 results in interaction with CTLA4 inducing an immunosuppressive tumor response.

The first interaction factor identified in *U1*-mutated samples involves a higher dependency of CD44 signaling through multiple targets, with the MIF-CD44-CD74 signaling particularly noteworthy. This interaction is described as promoting B-cell survival through the activation of the NFKB pathway^35^, consistent with our findings. This pathway has been extensively characterized for its anti-apoptotic effects in CLL, enhancing malignant B cell survival, as well as bone marrow homing, and resistance^36^. The hyperactivation of CD44 signaling might result from the overexpression and CD44v alternatively spliced isoforms in *U1*-mutated CLL cells^12^. These alterations, particularly CD44v6 generation, create a stop signal for circulating CLL cells via hyaluronan binding^37^, enabling lymph node retention and fostering proliferation. While this isoform is typically induced during CLL activation, it is constitutively expressed in 4% cases^38^. This subset aligns with the prevalence of the U1 g.3A>C mutation and exhibits significant overlap in an independent cohort, suggesting a potential mechanistic link. Beyond the well-characterized hyaluronan binding, cell-to-cell interaction simulation suggests that CD44 in *U1* mutated samples may exert its effects from systemic remodeling of extracellular matrix components, rather than specific molecular interactions. Regarding *SF3B1*-mutated samples, none of the identified interaction factors exhibited specificity, indicating that *U1* and *SF3B1* mutations elicit distinct responses within the CLL tumor microenvironment, despite converging on the same biological process.

Our analysis of the CLL microenvironment has provided a first glimpse at the complexity of interactions between the TME and CLL cells for cases with mutations in splicing related genes. The identified intercellular communication network offers promising avenues for alternative therapeutic strategies for U-CLL with mutations in *U1* snRNA. Thus, anti-CTLA-4 therapy^39^ could represent a novel approach due to the CD80/CD86 overexpression observed in *U1*-mutant samples. Notably, CD44, a key mediator of microenvironmental communication and intracellular signaling, was upregulated and alternatively spliced in *U1*-mutant CLL, positioning it as a promising target for therapies such as anti-CD44v6 treatment^40–42^. Together, these results provide an extensive characterization of the potentially critical influence of *U1* mutations on U-CLL development and identified CD44 as a key molecular player in this process.

## Methods

### Sample collection

A total of 26 tumor samples from 23 U-CLL patients from the CLL-ES study within the International Cancer Genome Consortium (ICGC) were used based on the mutational status of the splicing factor genes to be analyzed^6,8,13^. Bone marrow (n=15) and/or lymph node (n=11) biopsies were cryopreserved and stored in the IDIBAPS biobank. The cohort comprised 7 patients with the g.3A>C mutation in *U1 snRNA*, 6 with recurrent mutations in the *SB3F1* gene, predominantly the K700 and K666 variants, and as a control group, 10 U-CLL patients without mutations in those genes. Clinical and molecular characteristics of the included patients are shown in **Supplemental Table 1**.

### Sample processing

Cryopreserved bone marrow and lymph node samples were initially thawed in 5 ml of Iscove’s Modified Dulbecco’s Media (IDMD, Invitrogen) and centrifuged at 250 x g. The resulting pellets were washed twice with 2 ml of PBS. Then, the pellet was resuspended in 1 ml of PBS containing 0.04% bovine serum albumin (BSA), filtered through a 40 µm FlowMi cell strainer (Sigma) and diluted at a concentration between 700-1,000 cells/μl. Cell viability was confirmed to be >80% in all samples using 0.4% Trypan Blue dye with Countess 3 Automated Cell Counter (ThermoFisher).

### TME enrichment

To explore the cellular complexity of the tumor microenvironment (TME), 22 samples were analyzed directly without additional separation steps. However, for samples with a high tumor purity (>90%), measured at diagnosis by flow cytometry with the CD19 B-cell marker, we performed a TME enrichment step by depleting B-cells using the human CD19 Microbeads Kit and LD columns (Miltenyi Biotec) according to manufacturer’s protocol, employing the QuadroMACS™ Separator and the magnetic MACS® MultiStand (Miltenyi Biotec). The CD19 negative eluted fraction followed the sample preparation procedure mentioned above.

### Single-cell gene expression

For an estimated target between 5,000-10,000 cells, single-cell suspensions were loaded onto a Chromium Next GEM Chip K (10x Genomics) and processed using the iX Chromium (10x Genomics). Single-cell gel bead-in-emulsions were generated using the Chromium Next GEM Single Cell 5ʹ GEM kit v2, followed by a reverse transcription at 53 °C for 45 minutes in a Verity 96-well thermal cycler (Applied Biosystems). Single-strand cDNA, linked to a cell barcode and transcript unique molecular identifier (UMI), was purified by DynaBeads MyOne Silane Beads (Thermo Fisher Scientific) and amplified for 13 cycles (98 °C for 45 s; 98 °C for 20 s, 63 °C for 30 s, 72 °C for 1 min). cDNA quality was assessed with the TapeStation High Sensitivity D5000 ScreenTape (Agilent), and 50 ng were taken for the 5’ expression libraries with the library construction kit (10x Genomics). After 5 min of enzymatic fragmentation at 32°C, end-repair and A-tailing steps at 65°C for 30 min, followed by a 0.6-0.8X double size selection with SPRIselect beads (Beckman Coulter). The 50 µl eluate was adaptor ligated at 20°C for 15 min, and after post ligation cleanup step with 0.8X SPRIselect beads, a unique sample indexing PCR amplification was carried out using individual Dual Index TT Set A (10x Genomics) (98 °C for 45 s; 98 °C for 20 s, 54 °C for 30 s, and 72 °C for 20 s × 14 cycles; 72 °C for 1 min). Indexed libraries were subjected to a final 0.6-0.8X SPRIselect beads double-sided size selection. Single-cell RNA-seq library quality was confirmed with the TapeStation D1000 Screen Tape (Agilent) and the Qubit 3.0 dsDNA HS Assay Kit (Life Technologies).

### Single-cell TCR sequencing

To characterize T cell clonal expansion, T cell receptor (TCR) sequencing at single-cell resolution was performed, coupled with cell transcriptome in the CLL samples (n=23). Thus, 2 μl of the barcoded single-cell gene expression cDNA were used for V(D)J Amplification by the Single Cell Human TCR Amplification kit (10x Genomics). Full-length TCR V(D)J region was amplified through two enrichment steps with a heminested PCR strategy with V(D)J outer and inner primer pairs (98 °C for 45 s; 98 °C for 20 s, 62 °C for 30 s, and 72 °C for 1 min × 12 and 10 cycles; 72 °C for 1 min). A 0.5-0.8X double size selection using SPRIselect beads (Beckman Coulter) was later introduced. V(D)J amplicon quality was checked with the TapeStation D1000 Screen Tape (Agilent) and the Qubit 3.0 dsDNA HS Assay Kit (Life Technologies). Finally, 50 ng of the purified amplicons were taken for library generation following previously explained library construction kit instructions (10x Genomics).

### Single-cell surface protein

To improve manual annotation, a cohort of three CLL samples was incubated with the 5’ single-cell compatible TotalSeq™-C Human Universal Cocktail, V1.0 (BioLegend), specifically designed to profile the expression of 137 proteins at a single-cell level. Following manufacturer’s protocol, once the panel was reconstituted, cells were blocked with the Human TruStain FcX™ Fc Blocking reagent (BioLegend) and incubated for 10 minutes at 4°C. Later, the antibody cocktail was added and incubated for 30 min at 4°C, and washed twice with Cell Staining Buffer (BioLegend). Finally, stained cells were resuspended at a concentration of 1,000 cells/μl to proceed with the standard Chromium Next GEM Single Cell 5ʹ GEM kit v2 with Feature Barcode technology (10x Genomics).

### Sequencing

Equimolar amounts of indexed libraries were pooled and sequenced on an Illumina NovaSeq 6000 (Illumina). V(D)J and TotalSeq-C dual index libraries were sequenced at a minimum depth of 5,000 read pairs per/cell, while 5’ gene-expression dual index libraries were done at 25,000 read pairs per/cell. All libraries were sequenced using read lengths of 26 bp read 1, 10 bp i7 index, 90 bp read 2 and 10 bp i5 index. For long-read sequencing of *CD44* transcripts, 30 ng of the barcoded single-cell gene expression cDNA were used for targeted *CD44* amplification in two bone marrow samples with the *U1* g.3A>C mutation and two corresponding controls. Following SQK-PBK004 Nanopore protocol (Oxford Nanopore Technologies), *CD44* sequences were enriched by a four-primer PCR strategy, using custom FBP-forward primer: 5’-TTTCTGTTGGTGCTGATATTGCCTACACGACGCTCTTCCGA TCT-3’ and CD44 exon 17 reverse primer: 5’-ACTTGCCTGTCGCTCTATCTTCCTTCTTCGACTGGTGACTGCAA-3’, and individual indexes (98 °C for 3 min; 98 °C for 10 s, 68 °C for 20 s, and 72 °C for 20 s × 30 cycles; 72 °C for 2 min). Indexed libraries were later subjected to a final 0.65X SPRIselect beads (Beckman Coulter) left side size selection. Libraries quality was confirmed with the TapeStation D5000 Screen Tape (Agilent) and the Qubit 3.0 dsDNA HS Assay Kit (Life Technologies). 100 fmol of library pool was loaded on a Flo-Min114 MinION Flow Cell (Oxford Nanopore Technologies), and sequenced for 24 hours.

### scRNA-seq data processing

10x Genomics Cellranger software (6.1.2) was used to align, demultiplex and quantify unique molecular identifier (UMI) using the reference version hg38 (2020-A). Cellbender^43^ software (0.3.0) was deployed to diminish background noise and enhance the accuracy of gene expression estimation. Cellbender matrices were loaded into individual anndata objects using the Scanpy^44^ package (1.9.8). To filter cells, we used a two-step process. First, we applied a loose global filter, retaining only cells with more than 100 genes, more than 5% of ribosomal content and less than 20% of mitochondrial gene expression. Subsequently, we performed an intra-sample filter utilizing the median absolute deviation (MAD). By default, any cell deviating 5 MADs from the median in either direction was identified as an outlier, except for the mitochondrial percentage variable, filtered with 3 MADs. Doublets were detected by DoubletDetection(4.2)^45^. For all the downstream analyses, we considered the 10,000 most variable genes defined by highly variable function in Scanpy, with raw counts as input and flavor parameter fixed to “seurat_v3”.

Cell transcriptomes were embedded into a batch-corrected low-dimensional space using scVI^46^. Raw counts from the 10,000 most highly variable genes were used to train the scVI model. Throughout this paper, we used a total of 3 different models: one for all the cells, one for the tumor subset and another for T cells. The batch variable is the sequencing round, the primary covariables are the tissue and patient, and the epochs were determined for each model considering elbow validation parameters. We computed the UMAP embeddings for visualization using the latent space learned from scVI. Unsupervised clustering was also carried out using this learnt embedding with the Leiden algorithm. Diffusion map and pseudotime estimates were computed using the package PhenoGraph^47^ and Palantir^48^ for the CD8^+^ T cells subset. To infer transcription factor activities, we fit a univariate linear model within the decoupler framework^49^, based on the expression of downstream regulated genes. The activity, or score, was computed for each cell and defined as the t-score of the fitted model.

Cell type markers (**Supplemental Table 12** and **Supplemental Figure 1**), as well as Wilcoxon rank-sum differentially expressed genes across clusters were used for manual annotation of major cell types. T compartment cell subtypes were primarily defined through a label transfer approach using data from the study by Llaó et al^50^.

### Patient specificity

We calculated patient specificity scores using a shared nearest-neighbor graph. For each cell, patient specificity was determined by comparing the observed fraction of nearest neighbors to the expected fraction within the patient subgraph. The expected fraction of neighbors from the same patient was based on the overall fraction of cells for each patient. Patient specificity score was calculated as previously described.

### Expression gene signatures

For the identification of expression signatures, the data was extracted from the scVI batch-corrected latent space and normalized based on the following formula^17^:

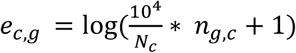

Where *c* refers to a specific cell and *g* to a gene. UMIs mapping to a gene *n_g,c_*are divided by the total number of counts *N_c_*. We removed immunoglobulin genes, as they are expected to have a very specific pattern in each patient, potentially masking other relevant transcriptional information. Additionally, we excluded mitochondrial, ribosomal and hemoglobin genes, as we observed their expression was mostly influenced by batch effects.

To decompose expression patterns and their condition-specific contribution, we used SignatureAnalyzer-GPU^51^, which employs a Bayesian adaptation of non-negative matrix factorization (ARD-NMF). Exponential priors were applied to both the *W’* and *H’* matrices, and a Gaussian distribution was assumed. The maximum number of iterations was fixed to 5,000 and the tolerance was left by default to 1 x 10^-5^. SignatureAnalyzer was run 10 times with random initialization for all the Ks between 1 and 25. We observed the elbow range between 7-15 (**Supplemental Figure 2A**). The solution with the minimum ARD objective function was selected as representative for each K value. To determine the optimal K parameter, hierarchical clustering of all programs within the predefined range was performed, revealing 12 robust metaprograms, akin to Gavish et al methodology^52^ (**Supplemental Figure 2B-D**). Consequently, we determined our optimum K to be 12.

To annotate the resulting patterns, we performed a gene-set enrichment analysis with the top rank genes in each pattern using KEGG and GO databases. To rank genes, we created a score that takes into account both weight and specificity as follows (**Supplemental Table 3**):

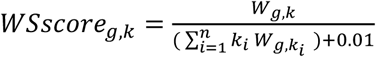

To determine if any of our patterns were correlated with our study variables, we performed both pairwise Spearman correlation and multivariate regression analyses for each pattern (**Supplemental Table 4**). A signature was labelled as “patient-specific” if its average activity across cells within any given patient exceeded 4 standard deviations compared to all other patients, and it was not considered for the downstream proportional and diversity analyses.

For differential expression analyses between conditions, we used a pseudobulk approach. For each patient, three pseudoreplicates were combined into distinct pseudobulk samples from the raw count data. PyDESeq2^53^ was employed to conduct differential expression analysis for each cell type, using Batch and Group as experimental design factors (**Supplemental Table 5**).

### Compositional analysis

We deployed the compositional analysis toolbox, scCODA^54^. A Bayesian generalized multivariate linear regression considering the tissue, batch and patient covariates was used. Cancer cell fraction from this analysis was removed to exclusively assess the relative relationships among the tumor microenvironment populations. The FDR was fixed at a conservative level of 0.2 according to the program specifications.

### TCR data analysis

The assembly and clonotype calling was performed using the Cellranger vdj software (v.6.1.2) against the prebuilt GRCh38 human reference (refdata-cellranger-vdj-GRCh38-alts-ensembl-7.0.0). Subsequent analyses were performed with the Python package Scirpy^55^ v.0.14.0. Productive TCRαβ chains were determined using the scirpy.tl.chain_qc function, and cells lacking V(D)J data or possessing two pairs of productive TCRαβ chains were excluded from the analysis. Patients with less than 100 cells with productive TCR were excluded from the subsequent analysis.

Clonotypes were determined based on the CDR3 nucleotide sequence identity and matching V-gene usage. Clonotype clusters were defined based on CDR3 amino acid pairwise sequence alignment with a cutoff= 10. Only clonotypes with more than two cells were considered. TCR clonality was assessed by computing Normalized Shannon Entropy and Diversity 50 (D50) index. Finally, TCR annotations were integrated with transcriptomic data to study clonality and TCR rearrangement at sub-celltype level.

### Ligand-receptor interaction analysis

Cell-to-cell interactions were quantified using LIANA^56^, integrating the CellPhoneDB, NATMI, and CellChat algorithms with *consensus* database as the reference. The aggregate rank from these algorithms’ output was computed using robust rank aggregation (RRA). We fixed the number of permutations at 100 and set the minimum number of cells per group to 10.

We inferred latent factors using a 4-dimensional NMF approach (TCA), as described in previous studies^57,58^. This allowed us to construct a tensor containing sample, sender cells, receiver cells, and ligand-receptor interactions information. We used the default workflow with the transformed magnitude score (1 - x) to make it suitable for the analysis. We performed the factorization fixing the “tf_optimization” parameter to robust, and to enhance reproducibility, we determined the initialization stage using Singular Value Decomposition (SVD). The optimal number of factors (K) was determined automatically comparing the decomposition outputs using CorrIndex, rather than classical elbow error metrics. Following the tutorial’s recommendation, we smoothed the curve using the Savitzky-Golay filter. This process identified 11 as the optimal number of factors (**Supplemental Figure 5A**).

### Statistical analysis

The analyses were conducted using multivariable multiple regression to assess the combined effects of tissue type and mutational status. For visualization purposes, pairwise significance shown in the figures was determined using the Mann-Whitney U test. Statistical significance was set at a threshold of *p*=0.05.

## Data and Code availability

The single-cell sequence datasets generated in this study are available for research purposes in the European Genome-phenome Archive (EGA) repository, under the accession number EGA50000000572. Additionally, the anndata, NMF, and TCA outputs can be accessed via Zenodo (DOI: 10.5281/zenodo.13835696). The code used in this paper is publicly accessible in a GitHub repository (https://github.com/xa-lab) for reproducibility.

## Data Availability

All data produced are available online at European Genome-phenome Archive (EGA) repository, under the accession number EGA50000000572. Additionally, the anndata, NMF, and TCA outputs can be accessed via Zenodo (DOI: 10.5281/zenodo.13835696). The code used in this paper is publicly accessible in a GitHub repository (https://github.com/xa-lab) for reproducibility.

## Acknowledgements

This study was supported by Ministerio de Ciencia e Innovación PID2020-117185RB-I00 and PID2023-148997OB-I00 to X.S.P., PID2021-123054OB-I00 to E.C. and PID2021-123165OB-I00 to D.C.; the Spanish Association Against Cancer (AECC PRYGN211258SUAR) to X.S.P.; “La Caixa” Foundation CLLSYSTEMS (HR22-00172); European Union NextGenerationEU/Mecanismo para la Recuperación y la Resilencia (MRR)/PRTR and the Instituto de Salud Carlos III (ISCIII) (PMP21/00015); Centro de Investigación Biomédica en Red Cáncer (CIBERONC). E.C. acknowledges the Generalitat de Catalunya Suport Grups de Recerca AGAUR (2021-SGR-01172). F.N. acknowledges research support from the American Association for Cancer Research (2021 AACR-Amgen Fellowship in Clinical/Translational Cancer Research, 21-40-11-NADE), European Hematology Association (EHA Junior Research Grant 2021, RG-202012-00245), and Lady Tata Memorial Trust (International Award for Research in Leukaemia 2021-2022, LADY_TATA_21_3223). E.C. is an Academia Researcher of the “Institució Catalana de Recerca i Estudis Avançats” (ICREA) of the Generalitat de Catalunya. S.L-T. is supported by a Severo Ochoa fellowship from the Asturian government. We also acknowledge support from the Institute of Oncology of Asturias (IUOPA, supported by Obra Social Cajastur, Spain).

## Author Contributions

E.C. and D.C. collected the samples and reviewed the pathology. J.F-M., S.L-T., C.O-F., A.A, processed the samples and generated the data by single-cell omics, S.L-T., J.F-M., P.B-M., X.S.P., designed the bioinformatics pipelines for the analysis of single-cell data. S.L-T., J.F-M., P.B-M., A.F., A.G-F, X.S.P. analyzed and interpreted the expression data. F.N, L.L-C., A.M.F., A.A., R.M-S., E.A.G, M.J., J.I.M-S. provided samples and/or data, and interpreted data. S.L-T., J.F-M., P.B-M., X.S.P., contributed to manuscript preparation. X.S.P. designed, reviewed and supervised the study. All authors reviewed, commented, and approved the final version of the manuscript.

