## Supplemental Figures for "*U1* snRNA mutation reshapes tumor microenvironment in chronic lymphocytic leukemia: a role for CD44-mediated signaling"

### Supplemental Figure Legends

**Supplemental Figure 1. Dataset description and marker gene expression.** (A) Barplot showing the number of cells after filtering per patient and eliminating cell doublets. (B-C) UMAP projection of scRNAseq data colored by tissue (B) and by condition(C). (D) Dot plot showing RNA marker gene expression for the major cell type classifications. Cell types are ordered by their frequency on the dataset. (E) Matrix plot showing mean CITE-seq markers expression for the major cell type classifications.

**Supplemental Figure 2. Analysis of tumor cell signatures.** (A) Boxplot showing the ARD objective function values for each run with different K parameter. The range highlighted in grey is the range used for hierarchical clustering and subsequent analyses. (B) UMAP projection of the different patterns obtained in the range between 7 and 15 colored by clustering. (C) Correlation heatmap and hierarchical clustering of all the patterns obtained in the range of 7 and 15. (D) Heatmap of the top genes within each pattern clustered by expression. (E) Signature weight per patient. Each cell was classified based on the signature by its highest score. (F) Dot plot showing the Gene Set Enrichment Analysis (GSEA) for the top 100 genes of the S7 signature. Boxplot showing the NFKB (G) and NFKBIB (H) activity inference based on target gene expression calculated with decoupler. (I) Dot plot showing the GSEA enrichment for the top 100 upregulated genes in U1 condition vs CTRL obtained from the pseudobulk differential expression analysis.

**Supplemental Figure 3. Analysis of the T cell compartment.** (A) Dot plot showing RNA marker gene expression for the T cell subpopulations. Cell types are ordered by their frequency on the dataset. (B) Matrix plot showing mean CITE-seq markers expression for the T cell subpopulations. (C) Enrichment of T cell type composition expressed in log2FC between lymph node and bone marrow. (D) Proportion of the different T cell subpopulations for each patient. (E) Gene expression dynamics of

CD8+T effector branch across pseudotime. Selected genes are labelled. (F) UMAP projection of T and NK compartment colored by pseudotime with the differentiation trajectory overlaid.

**Supplemental Figure 4. TCR and clonotype analysis.** (A) Bar plot with the total number of cells with productive TCR per patient after filtering. (B-C) Cumulative frequency of the top 10 clonotypes per patient grouped by tissue (B) and by condition (C). (D) Boxplot showing the median clone size per cell type and condition. (E-F) Stacked bar plot with the number of cells per cell type colored by clonotype expansion in control group (E) and in the *SF3B1* condition (F). (G) Number of clonotypes per cell type in the *U1* condition colored by expansion level. The shared clonotypes in more than five cells are highlighted in darker color. (H) Heatmap with pairwise Pearson correlation between D50 index and cumulative frequency and study variables in the CD8+ T cell compartment.

**Supplemental Figure 5. Cell to cell interaction analysis.** (A) Elbow plot showing (1- CorrIndex) and highlighting the optimal rank at 11. (B-C) Directionality of factor F10 (B) and factor F11 (C) specific interactions colored by cell type. (D-E) TCA factor weight per patient grouped by tissue (D) and condition (E). *, $p$<0.05. The p-values displayed in the figure correspond to pairwise comparisons and may differ from the multiple regression values shown in the table. (F) Boxplot showing the cellphonedb predicted strength of MIF>CD44^CD74 interaction between CLL cells grouped by condition. (G) Heatmap with pairwise Pearson correlation between CD44 expression and the most common genomic alterations in the studied cohort. (H-I) Boxplot showing the cellphonedb predicted strength of CD80 > CTLA4 interaction between CLL cells and $CD8^+$ T exhausted (H) and between CLL cells and $CD4^+$ Treg (I) grouped by condition.

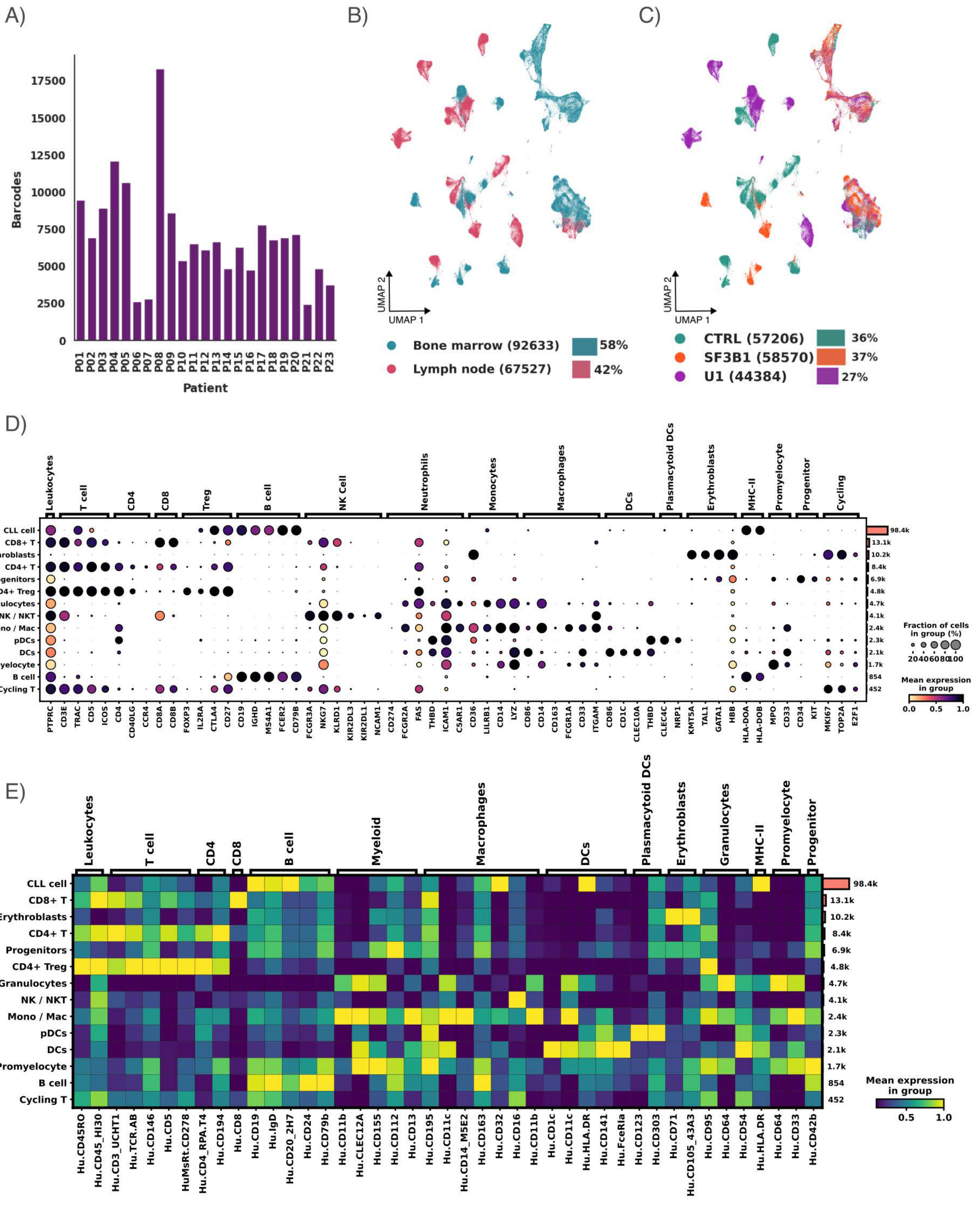

A)
Barcodes
0
2500
5000
7500
10000
12500
15000
17500
P01 P02 P03 P04 P05 P06 P07 P08 P09 P10 P11 P12 P13 P14 P15 P16 P17 P18 P19 P20 P21 P22 P23
Patient
B)
UMAP 2
UMAP 1
Bone marrow (92633) 58%
Lymph node (67527) 42%
C)
UMAP 2
UMAP 1
CTRL (57206) 36%
SF3B1 (58570) 37%
U1 (44384) 27%
D)
Leukocytes
T cell
CD4
CD8
Treg
B cell
NK Cell
Neutrophils
Monocytes
Macrophages
DCs
Plasmacytoid DCs
Erythroblasts
MHC-II
Promyelocyte
Progenitor
Cycling
CLL cell
CD8+ T
Erythroblasts
CD4+ T
Progenitors
CD4+ Treg
Granulocytes
NK / NKT
Mono / Mac
pDCs
DCs
Promyelocyte
B cell
Cycling T
98.4k
13.1k
10.2k
8.4k
6.9k
4.8k
4.7k
4.1k
2.4k
2.3k
2.1k
1.7k
854
452
PTPRC CD3E TRAC CD5 ICOS CD4 CD40LG CCR4 CD8A CD8B FOXP3 IL2RA CTLA4 CD27 CD19 IGHD MS4A1 FCER2 CD79B FCGR3A NKG7 KLRD1 KIR2DL3 KIR2DL1 NCAM1 CD274 FCGR2A FAS ICAM1 C5AR1 CD36 LILRB1 CD14 LYZ CD86 CD14 CD163 FCGR1A CD33 ITGAM CD86 CD1C CLEC10A THBD CLEC4C NRP1 KMT5A TAL1 GATA1 HBB HLA-DOA HLA-DOB MPO CD33 CD34 KIT MKI67 TOP2A E2F1
Fraction of cells in group (%)
20 40 60 80 100
Mean expression in group
0.0 0.5 1.0
E)
Leukocytes
T cell
CD4
CD8
B cell
Myeloid
Macrophages
DCs
Plasmacytoid DCs
Erythroblasts
Granulocytes
MHC-II
Promyelocyte
Progenitor
CLL cell
CD8+ T
Erythroblasts
CD4+ T
Progenitors
CD4+ Treg
Granulocytes
NK / NKT
Mono / Mac
pDCs
DCs
Promyelocyte
B cell
Cycling T
98.4k
13.1k
10.2k
8.4k
6.9k
4.8k
4.7k
4.1k
2.4k
2.3k
2.1k
1.7k
854
452
Hu.CD45RO Hu.CD45_HI30 Hu.CD3_UCHT1 Hu.TCR.AB Hu.CD146 Hu.CD5 HuMsRt.CD278 Hu.CD4_RPA.T4 Hu.CD194 Hu.CD8 Hu.CD19 Hu.IgD Hu.CD20_2H7 Hu.CD24 Hu.CD79b Hu.CD11b Hu.CLEC12A Hu.CD155 Hu.CD112 Hu.CD13 Hu.CD195 Hu.CD11c Hu.CD14_M5E2 Hu.CD163 Hu.CD32 Hu.CD16 Hu.CD11b Hu.CD1c Hu.CD11c Hu.HLA.DR Hu.CD141 Hu.FceRIa Hu.CD123 Hu.CD303 Hu.CD71 Hu.CD105_43A3 Hu.CD95 Hu.CD64 Hu.CD54 Hu.HLA.DR Hu.CD64 Hu.CD33 Hu.CD42b
Mean expression in group
0.5 1.0


Supp. Fig. 1

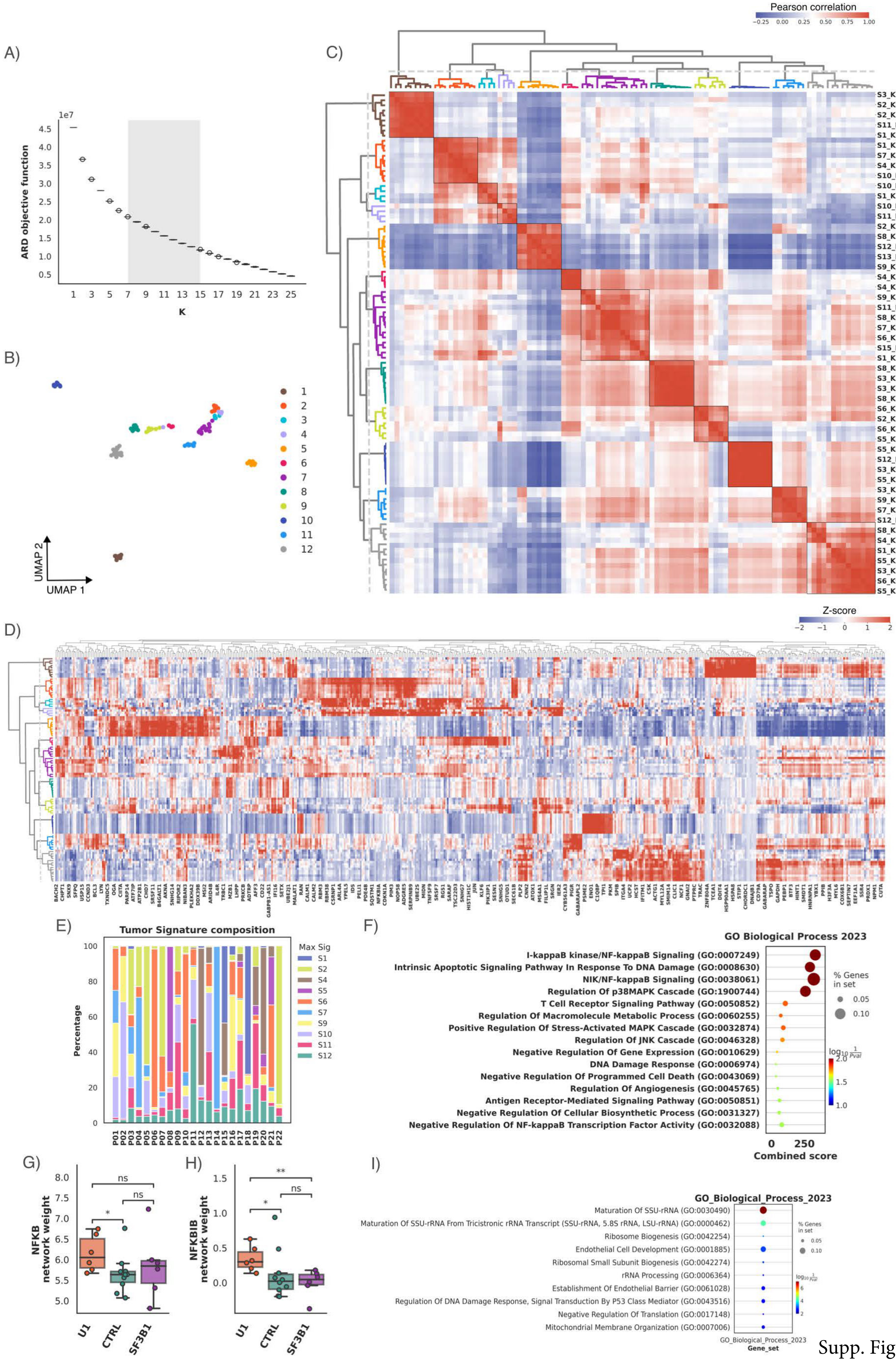
A)
ARD objective function
1e7
K
B)
UMAP 1
UMAP 2
C)
Pearson correlation
D)
Z-score
E)
Tumor Signature composition
Percentage
Max Sig
F)
GO Biological Process 2023
I-kappaB kinase/NF-kappaB Signaling (GO:0007249)
Intrinsic Apoptotic Signaling Pathway In Response To DNA Damage (GO:0008630)
NIK/NF-kappaB Signaling (GO:0038061)
Regulation Of p38MAPK Cascade (GO:1900744)
T Cell Receptor Signaling Pathway (GO:0050852)
Regulation Of Macromolecule Metabolic Process (GO:0060255)
Positive Regulation Of Stress-Activated MAPK Cascade (GO:0032874)
Regulation Of JNK Cascade (GO:0046328)
Negative Regulation Of Gene Expression (GO:0010629)
DNA Damage Response (GO:0006974)
Negative Regulation Of Programmed Cell Death (GO:0043069)
Regulation Of Angiogenesis (GO:0045765)
Antigen Receptor-Mediated Signaling Pathway (GO:0050851)
Negative Regulation Of Cellular Biosynthetic Process (GO:0031327)
Negative Regulation Of NF-kappaB Transcription Factor Activity (GO:0032088)
Combined score
% Genes in set
G)
NFKB network weight
U1
CTRL
SF3B1
H)
NFKBIB network weight
I)
GO_Biological_Process_2023
Maturation Of SSU-rRNA (GO:0030490)
Maturation Of SSU-rRNA From Tricistronic rRNA Transcript (SSU-rRNA, 5.8S rRNA, LSU-rRNA) (GO:0000462)
Ribosome Biogenesis (GO:0042254)
Endothelial Cell Development (GO:0001885)
Ribosomal Small Subunit Biogenesis (GO:0042274)
rRNA Processing (GO:0006364)
Establishment Of Endothelial Barrier (GO:0061028)
Regulation Of DNA Damage Response, Signal Transduction By P53 Class Mediator (GO:0043516)
Negative Regulation Of Translation (GO:0017148)
Mitochondrial Membrane Organization (GO:0007006)
Gene_set

Supp. Fig. 2

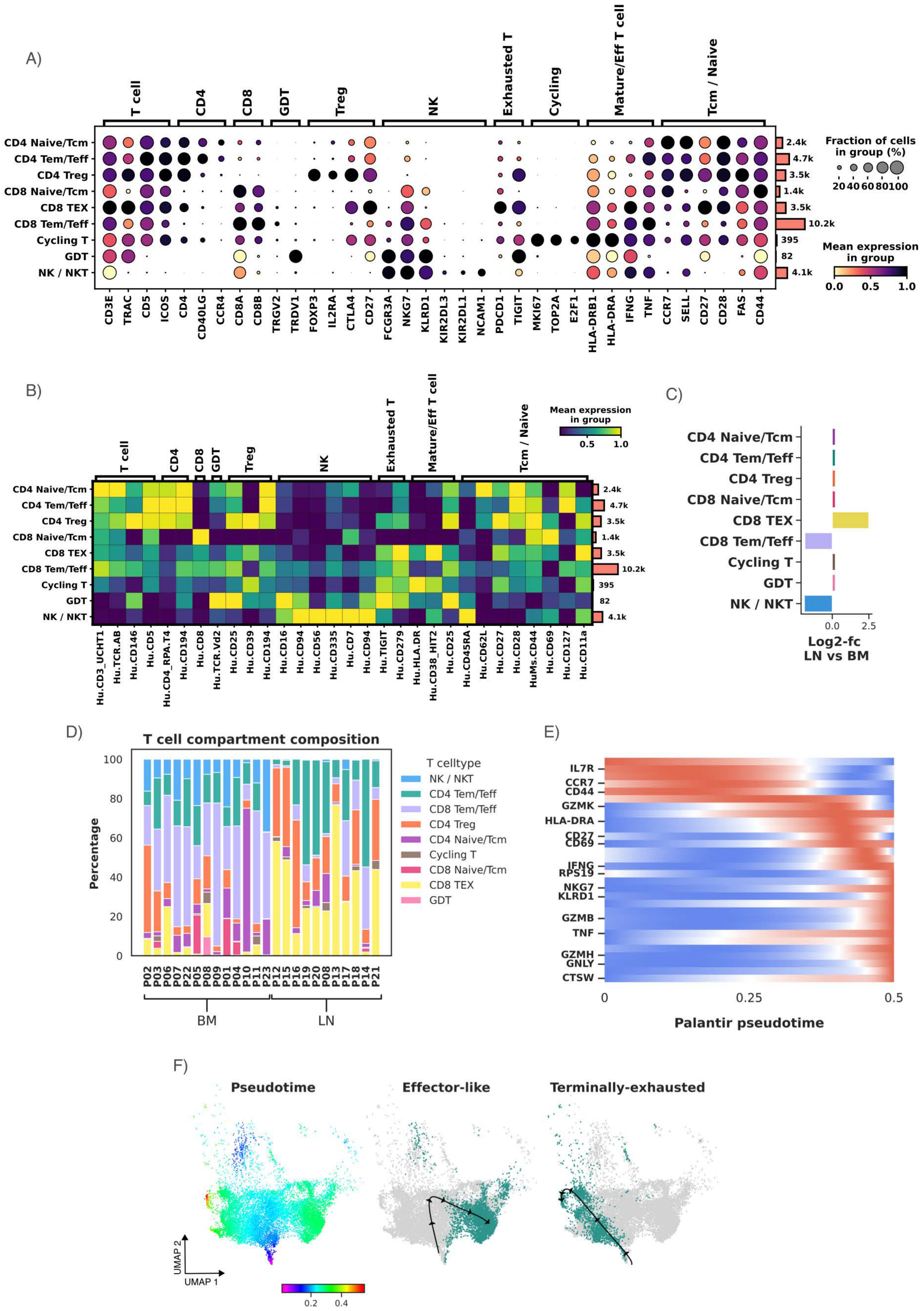
A)
T cell
CD4
CD8
GDT
Treg
NK
Exhausted T
Cycling
Mature/Eff T cell
Tcm / Naive
CD4 Naive/Tcm
CD4 Tem/Teff
CD4 Treg
CD8 Naive/Tcm
CD8 TEX
CD8 Tem/Teff
Cycling T
GDT
NK / NKT
2.4k
4.7k
3.5k
1.4k
3.5k
10.2k
395
82
4.1k
CD3E TRAC CD5 ICOS CD4 CD40LG CCR4 CD8A CD8B TRGV2 TRDV1 FOXP3 IL2RA CTLA4 CD27 FCGR3A NKG7 KLRD1 KIR2DL3 KIR2DL1 NCAM1 PDCD1 TIGIT MKI67 TOP2A E2F1 HLA-DRB1 HLA-DRA IFNG TNF CCR7 SELL CD27 CD28 FAS CD44
Fraction of cells in group (%)
20 40 60 80 100
Mean expression in group
0.0 0.5 1.0
B)
T cell
CD4
CD8
GDT
Treg
NK
Exhausted T
Mature/Eff T cell
Tcm / Naive
Mean expression in group
0.5 1.0
CD4 Naive/Tcm
CD4 Tem/Teff
CD4 Treg
CD8 Naive/Tcm
CD8 TEX
CD8 Tem/Teff
Cycling T
GDT
NK / NKT
2.4k
4.7k
3.5k
1.4k
3.5k
10.2k
395
82
4.1k
Hu.CD3_UCHT1 Hu.TCR.AB Hu.CD146 Hu.CD5 Hu.CD4_RPA.T4 Hu.CD194 Hu.CD8 Hu.TCR.Vd2 Hu.CD25 Hu.CD39 Hu.CD194 Hu.CD16 Hu.CD94 Hu.CD56 Hu.CD335 Hu.CD7 Hu.CD94 Hu.TIGIT Hu.CD279 Hu.HLA.DR Hu.CD38_HIT2 Hu.CD25 Hu.CD45RA Hu.CD62L Hu.CD27 Hu.CD28 HuMs.CD44 Hu.CD69 Hu.CD127 Hu.CD11a
C)
CD4 Naive/Tcm
CD4 Tem/Teff
CD4 Treg
CD8 Naive/Tcm
CD8 TEX
CD8 Tem/Teff
Cycling T
GDT
NK / NKT
0.0 2.5
Log2-fc LN vs BM
D)
T cell compartment composition
Percentage
0 20 40 60 80 100
P02 P03 P06 P07 P22 P05 P08 P09 P01 P04 P10 P11 P23 P12 P15 P16 P19 P20 P08 P13 P17 P18 P14 P21
BM
LN
T celltype
NK / NKT
CD4 Tem/Teff
CD8 Tem/Teff
CD4 Treg
CD4 Naive/Tcm
Cycling T
CD8 Naive/Tcm
CD8 TEX
GDT
E)
IL7R
CCR7
CD44
GZMK
HLA-DRA
CD27
CD69
IFNG
RPS19
NKG7
KLRD1
GZMB
TNF
GZMH
GNLY
CTSW
0 0.25 0.5
Palantir pseudotime
F)
Pseudotime
Effector-like
Terminally-exhausted
UMAP 2
UMAP 1
0.2 0.4

Supp. Fig. 3

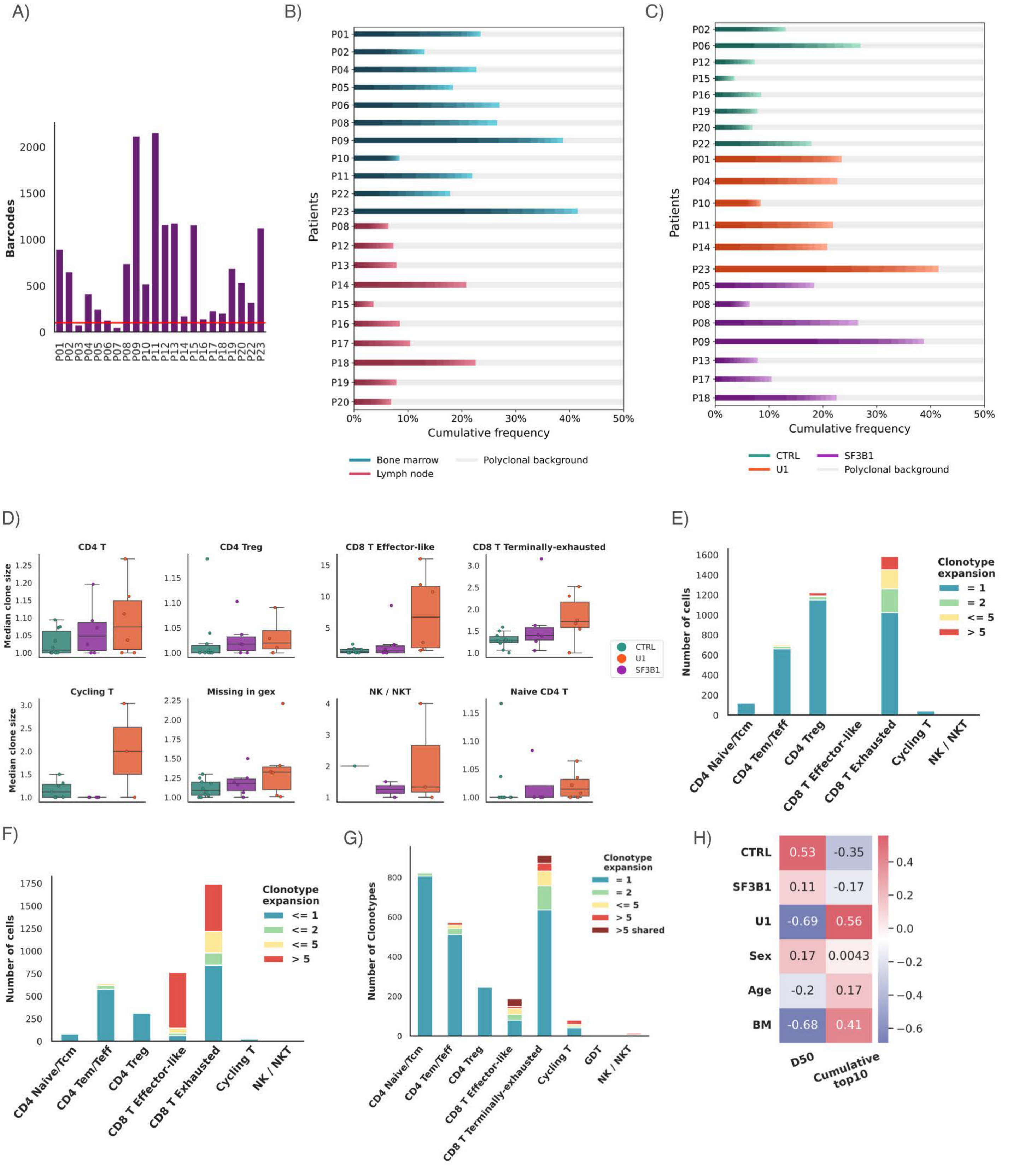
A)
Barcodes
B)
Patients
Cumulative frequency
Bone marrow
Lymph node
Polyclonal background
C)
Patients
Cumulative frequency
CTRL
U1
SF3B1
Polyclonal background
D)
CD4 T
CD4 Treg
CD8 T Effector-like
CD8 T Terminally-exhausted
Cycling T
Missing in gex
NK / NKT
Naive CD4 T
Median clone size
CTRL
U1
SF3B1
E)
Number of cells
Clonotype expansion
= 1
= 2
<= 5
> 5
CD4 Naive/Tcm
CD4 Tem/Teff
CD4 Treg
CD8 T Effector-like
CD8 T Exhausted
Cycling T
NK / NKT
F)
Number of cells
Clonotype expansion
<= 1
<= 2
<= 5
> 5
G)
Number of Clonotypes
Clonotype expansion
= 1
= 2
<= 5
> 5
>5 shared
CD8 T Terminally-exhausted
GDT
H)
CTRL
SF3B1
U1
Sex
Age
BM
D50
Cumulative top10

Supp. Fig. 4

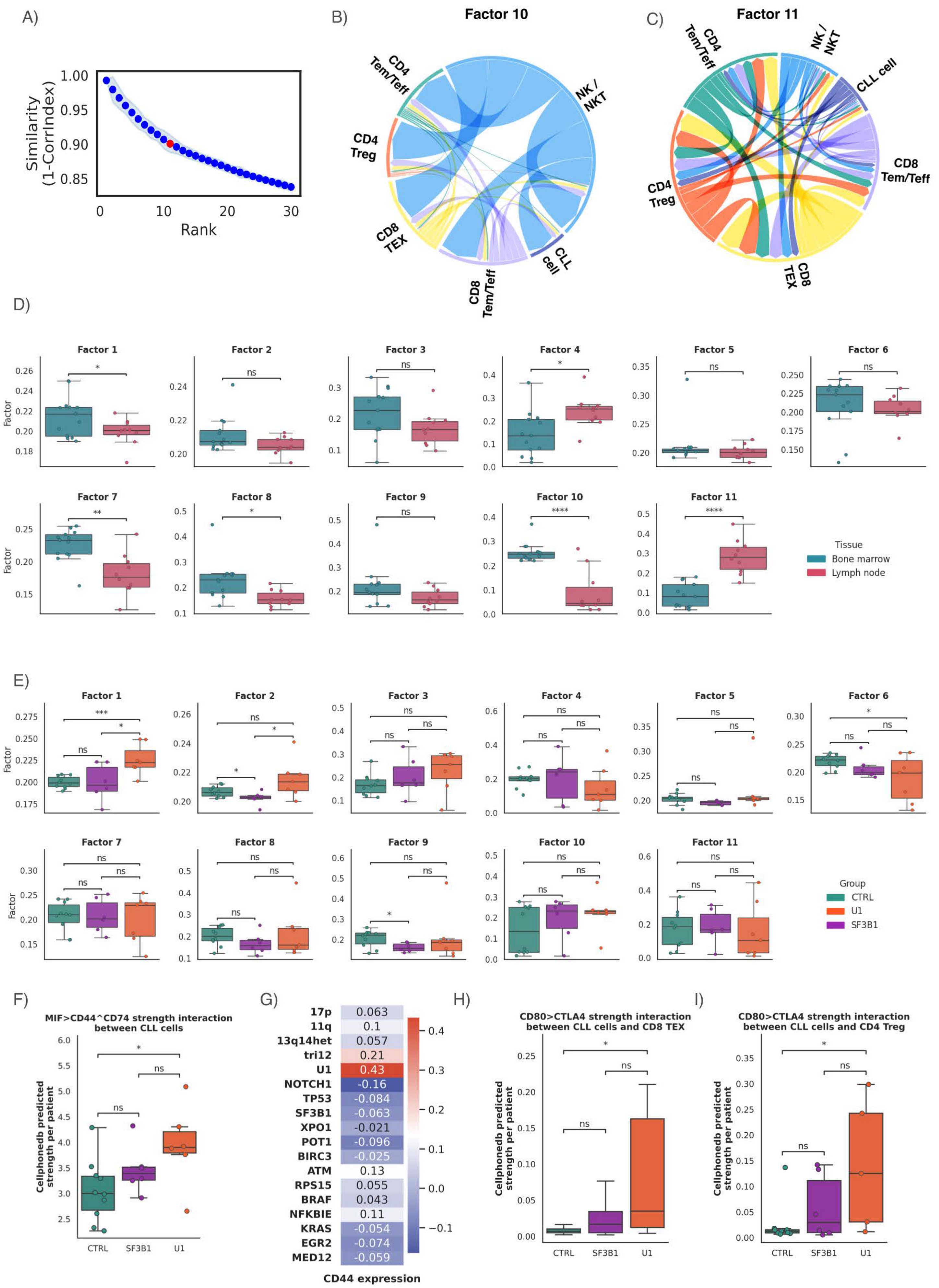
A)
Similarity (1-CorrIndex)
Rank
B)
Factor 10
CD4 Tem/Teff
NK / NKT
CD4 Treg
CD8 TEX
CD8 Tem/Teff
CLL cell
C)
Factor 11
CD4 Tem/Teff
NK / NKT
CLL cell
CD8 Tem/Teff
CD4 Treg
CD8 TEX
D)
Factor 1
Factor 2
Factor 3
Factor 4
Factor 5
Factor 6
Factor 7
Factor 8
Factor 9
Factor 10
Factor 11
Factor
Tissue
Bone marrow
Lymph node
E)
Group
CTRL
U1
SF3B1
F)
MIF>CD44^CD74 strength interaction between CLL cells
Cellphonedb predicted strength per patient
CTRL
SF3B1
U1
G)
17p 0.063
11q 0.1
13q14het 0.057
tri12 0.21
U1 0.43
NOTCH1 -0.16
TP53 -0.084
SF3B1 -0.063
XPO1 -0.021
POT1 -0.096
BIRC3 -0.025
ATM 0.13
RPS15 0.055
BRAF 0.043
NFKBIE 0.11
KRAS -0.054
EGR2 -0.074
MED12 -0.059
CD44 expression
H)
CD80>CTLA4 strength interaction between CLL cells and CD8 TEX
Cellphonedb predicted strength per patient
I)
CD80>CTLA4 strength interaction between CLL cells and CD4 Treg
Cellphonedb predicted strength per patient

Supp. Fig. 5
